# SARS-CoV-2 Seroprevalence Among Antenatal Clinic Attendees in Kingston, Jamaica, September-November 2020

**DOI:** 10.1101/2021.02.08.21251367

**Authors:** Te’Anne Chisolm, Jerome P. Walker, Ynolde Leys, Tiffany R. Butterfield, Candice Medley, Tamara K. Thompson, Glendee Reynolds-Campbell, Willem H. Mulder, Michelle Campbell Mitchell, Joshua J. Anzinger

**Affiliations:** The University of the West Indies, Department of Microbiology, Kingston, Jamaica, West Indies; The University of the West Indies, Department of Obstetrics and Gynaecology, Kingston, Jamaica, West Indies; The University of the West Indies, Department of Medicine, Kingston, Jamaica, West Indies; The University of the West Indies, Department of Chemistry, Kingston, Jamaica, West Indies

**Keywords:** SARS-CoV-2, seroprevalence, antibody test, pregnancy, Jamaica

## Abstract

SARS-CoV-2 seroprevalence in an antenatal population in Kingston, Jamaica was assessed for September-November 2020 in a repeated cross-sectional study using the Abbott Architect SARS-CoV-2 IgG assay. After adjusting for test performance, seroprevalence was 6.9% for September, 16.9% for October, and 24.0% for November. Of the 37 pregnant women testing SARS-CoV-2 IgG positive, only 3 were symptomatic. One symptomatic woman testing SARS-CoV-2 IgG positive had multiple co-morbidities and succumbed to COVID-19 pneumonia. Up to January 31, 2021, 8 women identified as SARS-CoV-2 IgG positive delivered, all without complications. Comparison of test adjusted seroprevalence data with cumulative PCR-confirmed COVID-19 cases within the Kingston Metropolitan Area indicated that as many as 44.4 times more people were infected with SARS-CoV-2 than identified with PCR testing. These findings provide the first evidence for the extent of SARS-CoV-2 infections in Jamaica and will inform future SARS-CoV-2 testing strategies.

---

The first confirmed COVID-19 cases in the Caribbean occurred in March 2020, with the first confirmed case of COVID-19 in Jamaica occurring on March 10, 2020.^1^ Subsequent to the introduction of SARS-CoV-2 in Jamaica, governmental restrictions were imposed that included school closures (March 13, 2020), closure of international borders (March 21, 2020), and implementation of daily island-wide curfews (April 1, 2020). Restrictions were eased on June 1, 2020 to reopen international borders, but up to January 2021 most schools have remained closed (virtual schooling) and island-wide curfews remain in effect. Up to August 2020, confirmed cases of COVID-19 in Jamaica remained below 1,000 and only 10 deaths were reported for the entire population of approximately 2.7 million people. Several weeks after the Emancipation Day (August 1, 2020) and Independence Day (August 6, 2020) holidays COVID-19 PCR-confirmed cases increased rapidly, followed by increased COVID-19 deaths, and on August 30, 2020 the Government of Jamaica declared COVID-19 community transmission. Up to January 31, 2021 there have been 15,973 confirmed COVID-19 cases and 353 deaths in Jamaica.

Although it is clear that SARS-CoV-2 community transmission in Jamaica has led to a great increase in COVID-19 cases and deaths, it remains unknown as to the extent of transmission, as even in the most resourced countries most cases are not identified.^2,3^ SARS-CoV-2 seroprevalence studies can determine the extent of transmission within a population that can inform the public health response. This information can indicate whether the amount of testing is adequate and also informs transmission dynamics due to persons likely having some degree of immunity that have been infected recently.^4^

Samples collected from pregnant women seeking routine antenatal care are commonly used to provide prevalence estimates of disease, as is done in some sub-Saharan African countries to determine HIV prevalence.^5^ More recently antenatal samples have been examined to provide a prevalence estimate of SARS-CoV-2 infections.^6^ Residual serum samples from pregnant women attending antenatal clinics provide a valuable tool to determine the extent of SARS-CoV-2 infections in a population and are also a unique population due to possible risks of infectious disease not only to the pregnant woman but also to the fetus. Although SARS-CoV-2-infected pregnant women are commonly asymptomatic,^7–9^ symptomatic pregnant women were recently shown to be at increased risk of severe disease.^10^ Thus far, studies of COVID-19 in pregnancy show that poor outcomes are uncommon for both the mother and child, and vertical transmission appears to be rare.^11^ COVID-19 studies in antenatal populations are limited, however, which has led several experts to promote increased COVID-19 surveillance and research of pregnant women.^12^

In this repeated cross-sectional study, we determined the presence of SARS-CoV-2 IgG in all University Hospital of the West Indies (UHWI) antenatal residual serum samples submitted during September-November 2020. The UHWI, located in Kingston, Jamaica, is the largest tertiary hospital in the country with approximately 1,600 deliveries each year. Most pregnant women attending the UHWI antenatal clinic reside in the Kingston Metropolitan Area, the most populous metropolitan area of Jamaica that is primarily within the parishes of Kingston and St Andrew. UHWI antenatal samples are routinely received in the Virology Laboratory of the University of the West Indies Department of Microbiology for HIV testing. A total of 249 unique patient samples (i.e., no patient was tested more than once) were tested for SARS-CoV-2 IgG. This study was approved by the UWI Mona Campus Research Ethics Committee (ECP 244 20/21).

SARS-CoV-2 IgG testing was determined via the Abbott chemiluminescence immunoassay (CMIA) using an Architect *i*2000SR instrument. The Abbott Architect SARS-CoV-2 IgG assay is CE marked and is EUA authorized by the FDA. Previous assessment of this assay in the adult Jamaican population did not include pregnant women in sensitivity analysis, but 20 samples from pregnant women were tested to determine specificity with one sample testing false positive (2.45 S/CO).^13^ An additional 32 antenatal samples collected in September-December 2019 (prior to SARS-CoV-2 introduction into Jamaica) were tested, with no samples testing SARS-CoV-2 IgG positive using the manufacturer’s recommended cutoff of ≥1.4 S/CO (Figure 1A). Including the previous data with this data resulted in a specificity of 98.08% (95% CI: 89.74-99.95%) when using the manufacturer’s recommended cutoff of ≥1.4 S/CO that maximizes specificity but underestimates true positives.^14,15^ Lowering the cutoff value from ≥1.4 S/CO to ≥0.4 S/CO resulted in one additional false positive sample (0.64 S/CO) for a specificity of 96.15% (95% CI: 86.79-99.53%). To determine the sensitivity of the Architect SARS-CoV-2 IgG assay for pregnant Jamaican women, we collected convalescent blood samples from SARS-CoV-2 PCR-confirmed pregnant women and tested sera for the presence of SARS-CoV-2 IgG (Figure 1B).Identification of SARS-CoV-2 PCR-confirmed pregnant women at the UHWI was challenging as most SARS-CoV-2 infected women were asymptomatic (see below). Of the 10 SARS-CoV-2 PCR confirmed pregnant women recruited, 8 tested positive (≥0.4 S/CO) for an overall sensitivity of 80% (95% CI: 44.39-97.48%).

**Figure 1.**
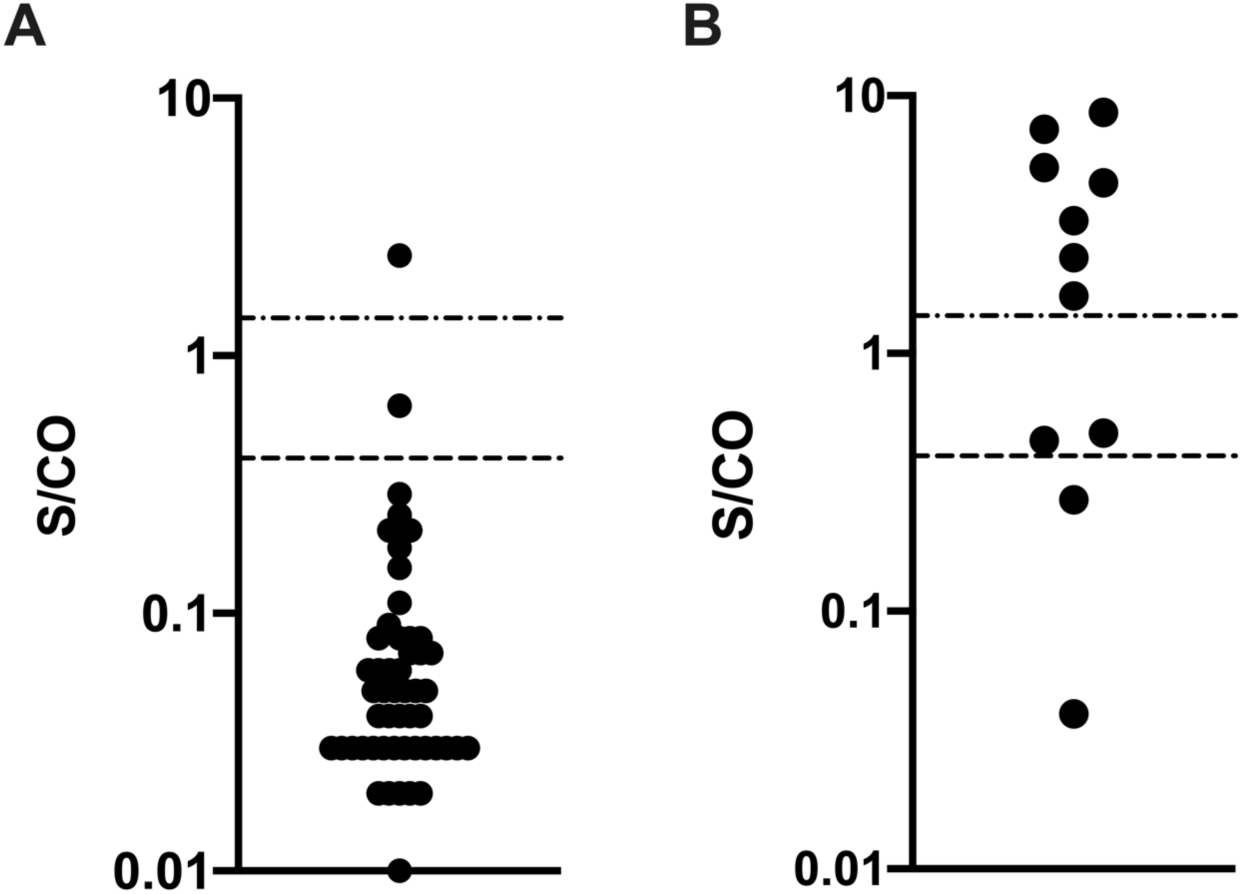
Architect SARS-CoV-2 IgG assay index values (S/CO) for (A) pre-pandemic 2019 antenatal samples and (B) convalescent sera from SARS-CoV-2 PCR confirmed pregnant women. The dashed lines indicate the manufacturer’s cutoff value (≥1.4 S/CO) and the cutoff value defined in this study for a positive test result (≥0.4 S/CO).

Samples from all women attending the UHWI antenatal clinic from September-November 2020 were tested, and 17 samples collected from the first week of March 2020 were also tested (Figure 2). The percentage of samples testing positive (≥0.4 S/CO) for each month was: 0.0% (0/17) in March, 9.1% (7/77) in September, 16.7% (13/78) in October, and 22.1% (17/77) in November (Table 1). Of the 37 women testing positive (≥0.4 S/CO), only 3 had a history of COVID-19 symptoms, consistent with previous observations that most pregnant women infected with SARS-CoV-2 remain asymptomatic.^7–9^ Only 8 of the 37 women testing positive (≥0.4 S/CO) had a SARS-CoV-2 PCR test up to the day of the sample collected that tested SARS-CoV-2 IgG positive, of which 3 were SARS-CoV-2 PCR positive. Of the 3 symptomatic women, 2 were SARS-CoV-2 PCR-confirmed, with 1 woman requiring intubation and management in the intensive care unit that subsequently demised. Of note the deceased patient was obese, with a history of breast cancer, and was of advanced maternal age (>40 years of age). Of the 8 mothers that delivered up to January 31, 2021, all neonates were born without complications with 2 deliveries occurring at late pre-term gestations. The distribution of ages was similar between women testing SARS-CoV-2 IgG negative and SARS-CoV-2 IgG positive as determined by Pearson’s Chi-squared test (*p*=0.164). Antenatal samples received for the months of September-November for 2018-2020 were similar (Pearson’s Chi-Square test; *p*=0.835), indicating that COVID-19 was unlikely to affect prenatal visit attendance during September-November 2020.

**Table 1.** Prevalence of SARS-CoV-2 infections in pregnant women.

| Month | Crude Prevalence<br>($\geq 1.4$ S/CO) | Crude Prevalence<br>($\geq 0.4$ S/CO) | Test Adjusted<br>Prevalence | Test and Seroreversion<br>Adjusted Prevalence |
| --- | --- | --- | --- | --- |
| March | 0.0% (0/17) | 0.0 (0/17) | — | — |
| September | 7.8% (6/77) | 9.1 (7/77) | 6.9 | 6.9 |
| October | 14.1% (11/78) | 16.7 (13/78) | 16.9 | 18.2 |
| November | 13.0 (10/77) | 22.1 (17/77) | 24.0 | 28.5 |

**Figure 2.**
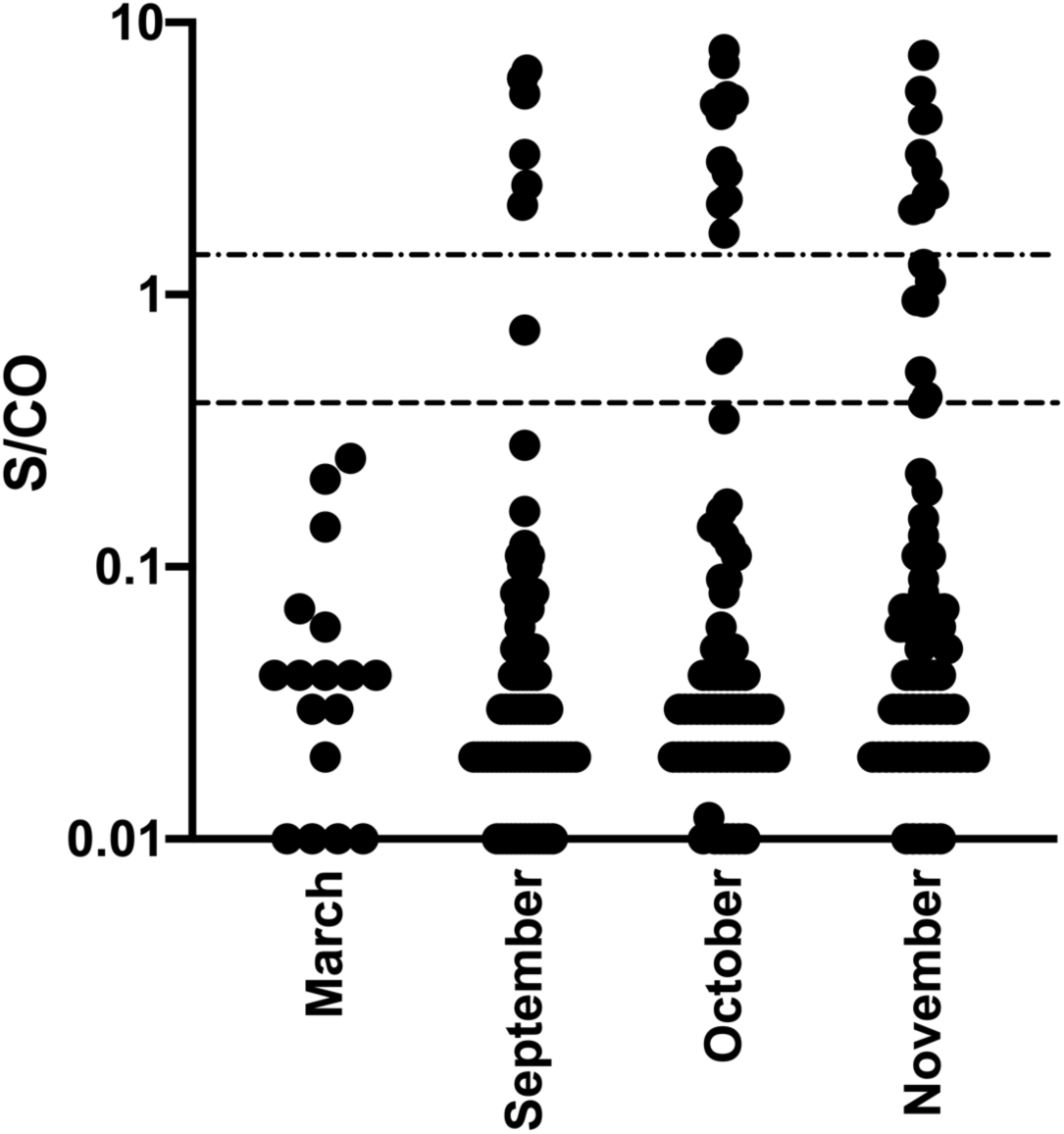
Distribution of SARS-CoV-2 IgG index values (S/CO) for tested residual antenatal serum samples. The dashed lines indicate the manufacturer’s cutoff value (≥1.4 S/CO) and the cutoff value defined in this study for a positive test result (≥0.4 S/CO).

For samples testing ≥0.4 S/CO, median monthly values showed a decreasing trend (Figure 2): 3.28 S/CO for September, 3.08 S/CO for October, and 2.08 S/CO for November (no samples in March tested ≥0.4 S/CO), indicating that seroreversion was likely occurring as has been reported previously for the Abbott Architect SARS-CoV-2 IgG assay.^15–18^ Crude seroprevalences were adjusted for test performance alone and with adjustment for seroreversion as described in the supplementary methods (Table 1). Adjusting for test performance only resulted in SARS-CoV-2 prevalences of 6.9% in September, 16.9% in October, and 24.0% in November. Adjusting for both test performance and seroreversion showed SARS-CoV-2 prevalences of 6.9% in September, 18.2% in October, and 28.5% in November.

Our data identifies an underappreciated prevalence of SARS-CoV-2 infections among antenatal women in the Kingston Metropolitan Area and provides an approximation of the extent of infections within the area. There were 2442, 3204, and 3590 cumulative confirmed COVID-19 cases in the parishes of Kingston and St Andrew in September, October, and November, respectively (Ministry of Health and Wellness). Thus, of the 669,773 persons inhabiting the parishes of Kingston and St Andrew (Statistical Institute of Jamaica, 2018), the percentage of the population identified to be infected with SARS-CoV-2 via PCR and serology (with test adjustment only), respectively, was 0.36% and 6.9% in September, 0.48% and 16.9% in October, 0.54% and 24.0% in November. This data indicates a 19.2-44.4-fold difference between serological identification of persons infected compared to SARS-CoV-2 PCR-confirmed cases, which is likely a slight underestimate due to seroreversion. The large disparity between SARS-CoV-2 PCR confirmed cases and those identified by antibody in this study highlights the difficulty of identifying SARS-CoV-2 cases with PCR testing, particularly in a resource-limited setting, and the utility of SARS-CoV-2 antibody testing to approximate population exposure.Although our study provides the first assessment of the extent of SARS-CoV-2 infections in Jamaica, SARS-CoV-2 has not spread throughout the island uniformly, limiting our ability to draw conclusions about the extent of virus infections throughout Jamaica. Future studies examining additional populations in Jamaica will be informative to identify the extent of SARS-CoV-2 circulation across the island.

## Data Availability

All data is included in the manuscript

## Acknowledgements

As a Global Infectious Diseases Scholars, Ynolde Leys and Tiffany Butterfield received mentored research training in the development of this manuscript. This training was supported in part by the University at Buffalo Clinical and Translational Science Institute award UL1TR001412 and the Global Infectious Diseases Research Training Program award D43TW010919. The content is solely the responsibility of the authors and does not necessarily represent the official views of the Clinical and Translational Science Institute or the National Institutes of Health.

## Financial Support

The University Hospital of the West Indies.

## Disclosure

All authors declared no conflict of interest.

## Authors’ addresses

Te’Anne Chisholm, Jerome Walker, Ynolde Leys, Tiffany Butterfield, Glendee Reynolds-Campbell, Joshua J. Anzinger, Department of Microbiology, The University of the West Indies, Mona, Kingston, Jamaica. Candice Medley, Michelle Campbell Mitchell, Department of Obstetrics and Gynaecology, The University of the West Indies, Mona, Kingston, Jamaica. Tamara K Thompson, Department of Medicine, The University of the West Indies, Kingston, Jamaica, Willem H Mulder, Department of Chemistry, The University of the West Indies, Kingston, Jamaica

## Notes

### Competing Interest Statement

The authors have declared no competing interest.

### Funding Statement

No external funding was received for this study.

### Author Declarations

UWI Mona Campus Research Ethics Committee (ECP 244 20/21)

