## Supplemental Methods for "SARS-CoV-2 Seroprevalence Among Antenatal Clinic Attendees in Kingston, Jamaica, September-November 2020"

**Prevalence adjusted for sensitivity and specificity of test**

Let $s$ = sensitivity = probability of true +, $f$ = probability of false +.
Then, specificity $\sigma = 1 - f$. Sensitivity- and specificity-adjusted prevalence = $t$.
Probability $p$ of + test result (= weighted prevalence):

$$p = \Pr(\text{true } +) + \Pr(\text{false } +) = ts + (1 - t)f.$$

Solve for $t$:

$$t = \frac{p - f}{s - f} = \frac{p + \sigma - 1}{s + \sigma - 1}.$$

In terms of percentages ($\rho = 100t\%$, $S = 100s\%$, $Sp = 100\sigma\%$, $\pi = 100p\%$),

$$\rho = \frac{\pi + Sp - 100\%}{S + Sp - 100\%} \times 100\%.$$

**Seroreversion-adjusted prevalence**

The vector $d[n] = \rho[n] - \rho[n-1] = r[n] - s[n]$, the difference between the fractions or %'s measured (*i.e.*, uncorrected for seroreversion) SARS-CoV-2 PCR confirmed *per capita* in months $n$ and $n - 1$ ($d[n]$ could be positive or negative), is related to the *actual* fraction or % new recoveries *per capita* in month $n$, $r[n]$, by the equation (Buss et al.)

$$d[n] = r[n] - \frac{p(1-\alpha)}{\alpha} \sum_{k=1}^{n-1} r[k]\alpha^{n-k},$$

or, in matrix notation, $\boldsymbol{d} = \boldsymbol{Ar}$, with the matrix elements of $\boldsymbol{A}$ given in terms of the model parameters $\alpha$ (decay rate, or monthly attenuation factor) and $p$ (proportion which sero-reverted) by

$$A_{i,j} = \begin{cases} 1 & i = j \\ -\dfrac{p(1-\alpha)}{\alpha}\alpha^{i-j} & i > j \\ 0 & i < j \end{cases}$$

($i, j = 1, \ldots, M$).
We are interested in the inverse relation, $\boldsymbol{r} = \boldsymbol{A}^{-1}\boldsymbol{d}$.
The (triangular) matrix $\boldsymbol{A}$ can be inverted. The elements of $\boldsymbol{A}^{-1}$ are found to be given by

$$(A^{-1})_{i,j} = \begin{cases} 1 & i=j \\ \dfrac{(\alpha + p(1-\alpha))^{i-j}}{1+\dfrac{\alpha}{p(1-\alpha)}} & i>j \\ 0 & i<j \end{cases}$$

Hence, the corrected prevalence *per capita* follows from the measured one as

$$r[n] = d[n] + \left(1+\frac{\alpha}{p(1-\alpha)}\right)^{-1} \sum_{k=1}^{n-1} d[k](\alpha + p(1-\alpha))^{n-k}.$$

The 'cost function' proposed by Buss et al. is defined as

$$J(\alpha, p) = r[M] + r[M-1]$$

and is to be minimised w.r.t. the two independent variables $\alpha$ and $p$, both of which are in the range [0, 1].
Substitution for the $r$ values from the previous equation yields

$$J(\alpha,p) = d[M] + d[M-1] + \left(1+\frac{\alpha}{p(1-\alpha)}\right)^{-1} \left[\sum_{k=1}^{M-1} d[k](\alpha+p(1-\alpha))^{M-k} + \sum_{k=1}^{M-2} d[k](\alpha+p(1-\alpha))^{M-k-1}\right].$$

We may now define two new independent variables,

$$x \equiv \alpha + p(1-\alpha) \text{ and } y \equiv \left(1+\frac{\alpha}{p(1-\alpha)}\right)^{-1},$$

and consider $J$ to be a function of $x$ and $y$ instead, and minimize it w.r.t. these new variables (which are also both in the range [0, 1]). Now, the square-bracket factor is a function of $x$ alone, call it $f(x)$, so $J$ is of the form

$$J(x, y) = \text{constant} + yf(x).$$

If there is a (global) minimum, it should occur where $\partial J/\partial x = 0$ and $\partial J/\partial y = 0$, producing two simultaneous equations in $x$ and $y$, the solution of which can then be transformed back to $\alpha$ and $p$. However, applying it to $J$ as given, these criteria lead to $f(x) = 0$ and $df(x)/dx = 0$ (since $y \neq 0$), which cannot be satisfied for any single value of $x$ (*i.e.* the problem is over-determined).
